# Remote Actigraphy Is Seasonally Variable with Implications for Clinical Monitoring

**DOI:** 10.1101/2025.05.22.25328110

**Authors:** Alexander M K Rothman, Jennifer Middleton, Hamza Zafar, Eckart M.D.D. De Bie, Joseph Newman, Frances Varian, Jake Taylor, Felicity Hitchcock, Cameron Ashraf, Junaid Patel, A. A. Roger Thompson, Mark Toshner

## Abstract

Remote monitoring technologies are increasingly used to assess physical activity and cardiopulmonary haemodynamics in patients with pulmonary arterial hypertension (PAH), yet the seasonal stability of these parameters remains poorly defined. In this study, patients with PAH underwent continuous remote monitoring using implantable cardiac monitors, pulmonary artery pressure sensors, smartwatches, and structured hospital-based walk tests to assess active and passive physical activity and haemodynamic parameters throughout the year. Passively collected physical activity data demonstrated significant seasonal variation, with the lowest levels in winter and peak activity during summer. In contrast, no seasonal variability was observed in structured exercise tests (incremental shuttle walk test, 6-minute walk test) or remote measures of pulmonary artery pressure, cardiac output, heart rate, or heart rate variability. Furthermore, national clinical trial recruitment data revealed seasonal fluctuations, with reduced enrolment in April and December. These findings underscore the importance of accounting for environmental and temporal factors when interpreting passively collected physical activity data and designing clinical trials that incorporate remote digital endpoints.

## Introduction

Remote measurement of physical activity and cardiopulmonary haemodynamics is increasingly being used in clinical practice and clinical trials.^1,2^ The FDA has accepted actigraphy as a primary endpoint for trials in interstitial lung disease-associated pulmonary hypertension,^3^ and the European Medicines Agency (EMA) has qualified the digital outcome measure Stride Velocity 95 Centile (SV95C) in 2023 as a primary endpoint for clinical trials of Duchenne muscular dystrophy.^4^ Actively generated activity data, such as a field-walk test, is an established clinical tool and regulatory endpoint, however, passively generated activity data, which may more closely reflect the activities undertaken by a patient in a real-world setting, remains investigational. The seasonal variation of active and passive physical activity measures, and cardiopulmonary haemodynamics has not been established.

We sought to evaluate the seasonal stability of hospital-based field-walk test, remote-monitored active and passive physical activity measures and remote-monitored haemodynamic parameters.

## Methods

Patients with a confirmed diagnosis of pulmonary arterial hypertension were enrolled in the UK National Cohort Study of Idiopathic and Heritable Pulmonary Arterial Hypertension (13/EE/0203). Two sub-studies of the National Cohort study were established for 1) implantation of an insertable cardiac monitor (ICM, LinQ, Medtronic). Physical activity was measured from a three-axis accelerometer within the ICM through a regulatory-approved online portal between November 2018 and October 2023.^5^ A subset of patients were co-enrolled in Feasibility of Novel Clinical Trial Infrastructure, Design and Technology for Early Phase Studies in Patients with Pulmonary Hypertension (FIT-PH, NCT04078243, REC 19/YH/0354) and a pulmonary artery pressure monitor (PAP monitor, CardioMEMS, Abbott) implanted; 2) provided participants with a smartwatch (Venu 2, Garmin) on their non-dominant wrist, paired with a customised digital walking module on the Atom5TM (Aparito Ltd) mobile application on the patients’ smartphone (iOS or Android).

Field-walk test: incremental shuttle walk test data from 5,820 consecutive patients were analysed from the ASPIRE registry (REC: 22/EE/0011) database between February 2001 and May 2019^5,^ and 1001 baseline 6MWT data from patients enrolled in the UK National Cohort Study of Idiopathic Pulmonary Arterial Hypertension between May 2014 and 2023. All patients had undergone systematic evaluation, including echocardiography, blood testing, exercise testing (incremental shuttle walk test), lung function testing, multimodality imaging,^6^ and right heart catheterisation, in accordance with nationally agreed and audited standards of care. No patients were lost to follow-up.

NIHR portfolio recruitment: De-identified summary statistics for all NIHR portfolio registered studies between January 2008 and December 2018 were identified through the NIHR Open Data Platform.

Statistics: parametric data is presented as mean +/−SEM, and non-parametric as median with interquartile range. Statistical significance was determined using one-way ANOVA or the Chi-square test as appropriate. Analysis was undertaken in Prism (10.0.3) for macOS.

## Results

In patients with an insertable cardiac monitor, average daily physical activity varied throughout the year, ranging from 129.5 min/day to 155.0 min/day (Figure 1A). Physical activity was cyclical and significantly altered by season, with physical activity most limited in winter (136 min ± 2.5 (SEM)), increasing through autumn (141min ± 4.9 (SEM)) and spring (149 min ± 2.6 (SEM)) and greatest in summer (151min ± 2.4 (SEM)) (Figure 1B). No seasonal variation was observed in ISWD or 6MWD (Figure 1C and D). Additionally, no seasonal variation was observed for heart rate, heart rate variability, pulmonary artery pressure or cardiac output (Figure 1A).

**Figure 1:**
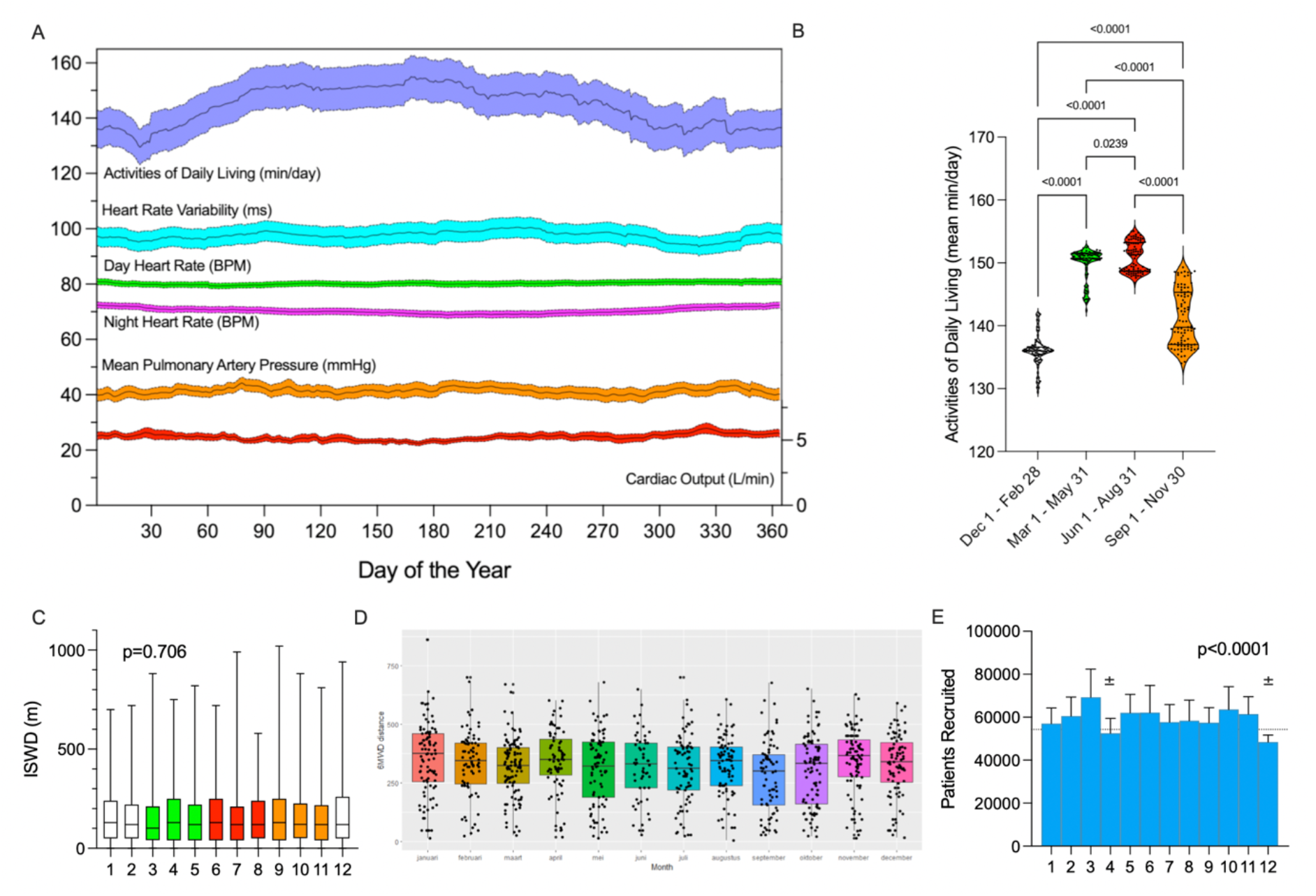
**A** - Physical activity (blue), heart rate variability (cyan), day heart rate (green), night heart rate (purple), mean pulmonary artery pressure (orange) and cardiac output (red) presented by day of the year (mean +/−SEM). **B** - Physical activity by calendar season (one-way ANOVA). **C-E** - Incremental shuttle walk test (C, n=5820), six-minute walk distance (D, n=1001) and NIHR portfolio studies (F, n=7,816,287) by calendar month (field-walk test one-way ANOVA, NIHR portfolio recruitment Chi-square).

To evaluate the seasonal variation of clinical study recruitment, we used data from 7,816,287 participants enrolled in UK NIHR portfolio clinical trials. The number of participants recruited was lower than expected in April and December (Figure 1D, p<0.0001).

## Discussion

The development of technology that provides high-dimensional physical activity and cardiopulmonary function data from a patient’s home offers the potential to revolutionise patient evaluation in clinical practice and research studies.^7^ The present study leveraged minimally invasive technology to evaluate the stability of physical activity, cardiopulmonary haemodynamics and hospital-measured field walk test through the calendar year. While active measures of physical activity and cardiopulmonary haemodynamics remained stable throughout the calendar year, passively collected physical activity showed a marked seasonal variability. The lack of variation in established measures of disease severity throughout the calendar year suggests that the observed variability of physical activity is attributed to changes in external environmental conditions, rather than a seasonal effect on disease status.^8^

The evaluation of remotely measured physical activity has already been incorporated into clinical trial designs due to the ease of measurement and its importance to patients and regulators indicated in patient surveys^9^ and FDA guidance.^10^ The observed variation in clinical trial recruitment and seasonal changes in physical activity introduce additional nondifferential errors, which may bias towards the null hypothesis. Understanding the seasonal variability of remote measures will facilitate the development of novel clinical study designs and appropriate data analysis. Given the global nature of trial recruitment and the likely variation in the impact of seasonality on activity and recruitment by geographic location, accounting for these biases in study design and analysis plans will not be simple. Seasonal variation in physical activity may also render data challenging to interpret and uninformative to clinical decision-making.

The high-dimensional data provided by home monitoring offers the potential to reduce signal-to-noise, provide a measurement more reflective of patients’ real-world experience and empower patients to have more direct ownership of the means of monitoring their disease. Beyond the fundamental question of how to best quantify activity, major challenges exist related to the standardisation of data processing and analyses. The present study highlights a major confounder that may further limit the interpretation of passively collected physical activity data when compared to active structured activity measurement in clinical trials and clinical practice.

## Conclusions

Seasonal variation is observed in remote actigraphy, but not remote-monitored cardiopulmonary physiology or structured exercise testing. Clinical trials, which also vary in their seasonal recruitment, will need to mitigate this confounding effect in design considerations. Seasonal changes in actigraphy are important as activity measures become more common in clinical care.

## Data Availability

All data produced in the present study are available upon reasonable request to the authors

